# The Prediction for Development of COVID-19 in Global Major Epidemic Areas Through Empirical Trends in China by Utilizing State Transition Matrix Model

**DOI:** 10.1101/2020.03.10.20033670

**Authors:** Zhong Zheng, Ke Wu, Zhixian Yao, Junhua Zheng, Jian Chen

## Abstract

**Background:** Since pneumonia caused by coronavirus disease 2019 (COVID-19) broke out in Wuhan, Hubei province, China, tremendous infected cases has risen all over the world attributed to high transmissibility. We managed to mathematically forecast the inflection point (IFP) of new cases in South Korea, Italy, and Iran, utilizing the transcendental model from Hubei and non-Hubei in China.

**Methods:** We extracted data from reports released by the National Health Commission of the People’s Republic of China (Dec 31, 2019 to Mar 5, 2020) and the World Health Organization (Jan 20, 2020 to Mar 5, 2020) as the training set to deduce the arrival of the IFP of new cases in Hubei and non-Hubei on subsequent days and the data from Mar 6 to Mar 9 as validation set. New close contacts, newly confirmed cases, cumulative confirmed cases, non-severe cases, severe cases, critical cases, cured cases, and death data were collected and analyzed. Using this state transition matrix model, the horizon of the IFP of time (the rate of new increment reaches zero) could be predicted in South Korean, Italy, and Iran. Also, through this model, the global trend of the epidemic will be decoded to allocate international medical resources better and instruct the strategy for quarantine.

**Results:** the optimistic scenario (non-Hubei model, daily increment rate of −3.87%), the relative pessimistic scenario (Hubei model, daily increment rate of −2.20%), and the relatively pessimistic scenario (adjustment, daily increment rate of −1.50%) were inferred and modeling from data in China. Matching and fitting with these scenarios, the IFP of time in South Korea would be Mar 6-Mar 12, Italy Mar 10-Mar 24, and Iran is Mar 10-Mar 24. The numbers of cumulative confirmed patients will reach approximately 20k in South Korea, 209k in Italy, and 226k in Iran under fitting scenarios, respectively. There should be room for improvement if these metrics continue to improve. In that case, the IFP will arrive earlier than our estimation. However, with the adoption of different diagnosis criteria, the variation of new cases could impose various influences in the predictive model. If that happens, the IFP of increment will be higher than predicted above.

**Conclusion:** We can affirm that the end of the burst of the epidemic is still inapproachable, and the number of confirmed cases is still escalating. With the augment of data, the world epidemic trend could be further predicted, and it is imperative to consummate the assignment of global medical resources to manipulate the development of COVID-19.

## Introduction

Since the first case of novel coronavirus pneumonia (NCP), caused by coronavirus disease 2019 (COVID-19), occurred in Wuhan, Hubei Province, China, the dreadful epidemic broke out during Dec 2019-Mar 2020 under the pace of Chinese Spring Festival [1]. With the untiring efforts of the people and the selfless dedication of medical staff, a total of 59,897 cured patients were discharged [2]. As of 24:00 on Mar 9, China has accumulated a total of 80,754 confirmed cases (including 4,794 severe cases) and 3136 dead cases [3]. However, in January, when the large-scale outbreak in China began, the disease initiated to spread to other parts of the world [4, 5]. As of Mar 9, a total of 7,382 cases were confirmed in South Korea, 7,375 cases in Italy, and 6,566 cases in Iran [6].

Similar to another coronavirus (CoV) —SARS-CoV— COVID-19 is an RNA virus that contains particular spike proteins conjugating with angiotensin-converting enzyme 2 (ACE2) that widely expressed in different tissues [7]. However, its doughty transmissibility in the community has a strong correlation with reasons such as long incubation period, mild early symptoms, and the like [8, 9]. Even though some studies have proved that Remdesivir designed for the Ebola virus may have a promising effect on COVID-19 [10], the kernel strategies for the prevention and treatment of NCP are still effective quarantine as in the case of SARS [11]. After the implementation of strict isolation, the most significant thing is the arrival of the peak and inflection point (IFP) of new cases of NCP for assessing the efficacy of current strategies.

Based on the previous data, we analyzed the epidemic situation in Hubei Province [12]. After the validation of the model, we were able to analyze the world epidemic trends, and predict the arrival of peaks and IFPs of newly confirmed cases and provide references for NCP prevention and control strategies in various countries.

## Method

### Study Population, Data Collection, and analysis

Data from reports, including medical observation, close contacts, confirmed cases, severe cases, critical cases, cured cases and death data and corresponding information, released by the Health Commission of Hubei Province (HCHP) (Dec 31, 2019 to Feb 8, 2020) were extracted as the training set. Primarily, the arrival of the IFP of new cases and epidemic trends in Hubei were deduced and testified in the validation set, whose data were extracted from HCHP (Feb 9, 2020 to Mar 5, 2020). Subsequently, another training set consisting of the data from the National Health Commission of the People’s Republic of China (NHC) and the World Health Organization (WHO) (Jan 20, 2020 to Mar 5, 2020) were established. Eventually, the data, including cumulative confirmed cases, cumulative cured cases, death data and corresponding information, from NHC and WHO (Mar 6, 2020 to Mar 9 2020) were collected and constructed the validation set. The data period starts from Dec 31, 2019 to Mar 9, 2020. Data is updated on the daily basis. All data were analyzed using Microsoft Excel (Microsoft Office 2016) and R studio (R Foundation for Statistical Computing, Vienna, Austria). The world epidemic situation was performed using the *nCov2019* package of R [13]. Histogram was obtained using the *ggplot2* packages of R.

### State Transition Matrix Model

State transition matrix (STM) modeling is a well-regarded approach widely applied in clinical decision analysis based on computer simulation. For estimating the IFP of newly confirmed cases and the scale of cumulative cases in the globe in subsequent days, we chose the Markov model cohort simulation.

### Parameter Selection and Estimate

In order to estimate the risk metrics (infectivity, severity, lethality) of the NCP, we build a STM model as showing in the figure (Figure.1).

**Figure 1.**
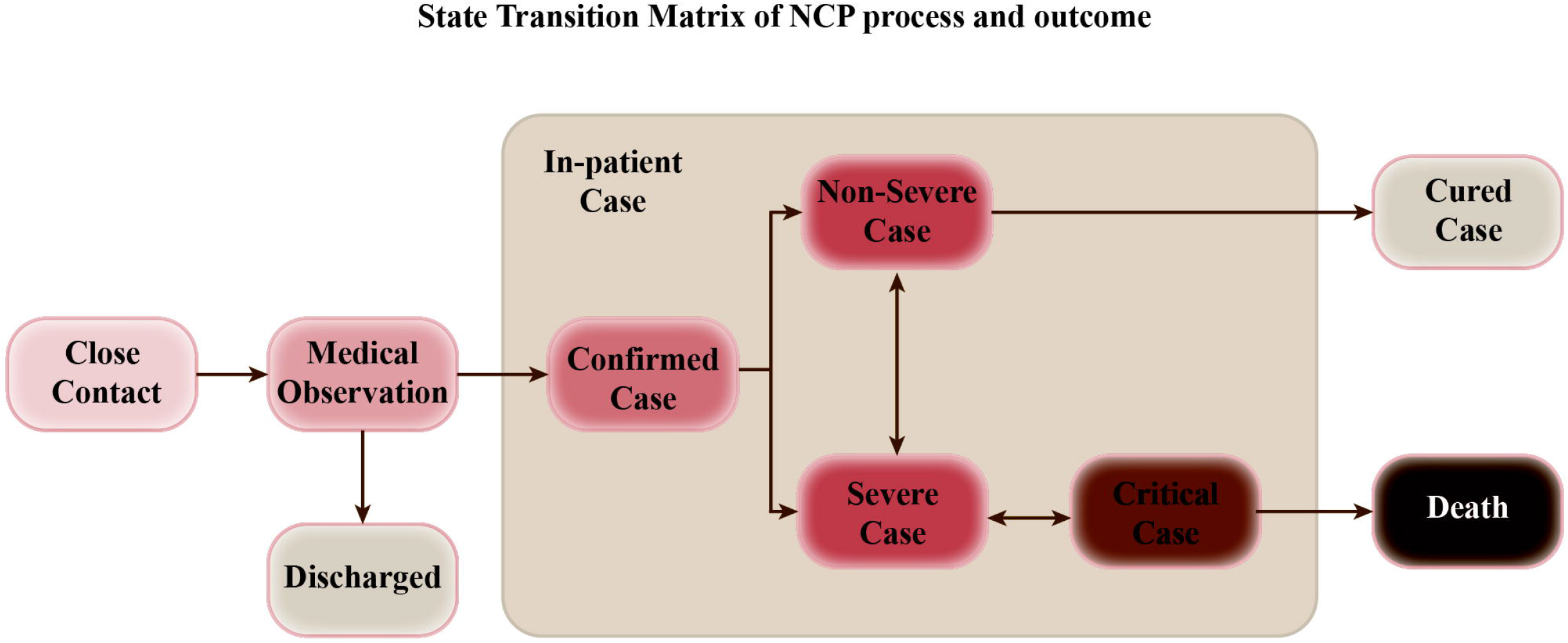
Process and outcome of the State Transition Matrix establishment when a Close Contact develops into the state of Medical Observation.

We define the states in this model. Medical Observation (MO) is the close contact of confirmed cases and put into medical observation. In the subsequent days, outcome could be any of the three: confirmed cases, discharged without COVID-19 infection, or stay in MO. Discharge (Disc) is a terminal state for a close contact, until he or she becomes another incident of close contact again. Infected is an intermediate state, where the patient becomes a confirmed infected case. The outcome is binary: severe, or non-severe. And the outcome is revealed immediately. Non-Severe Case (NS) is the patient also has three possible outcomes in the next day: cure, severe case, or stay in non-severe case. Severe Case (S), the patient has three possible outcomes in the next day: critical case, non-severe case, or stay in severe case. Critical Case (Cr), the patient has three possible outcomes in the next day: cured case, severe case, or stay in critical case. Cured Case (Cu) and Death (D) are also the terminal states for the patient. So, at any moment, we can identify the close contact or patient’s state by utilizing a state vector, defined as the following:

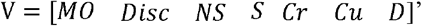

Where each element of the vector stands for one state in the same sequentially arranged order as mentioned above. Please note that the comfirmed itself is not an independent state, since the outcome is revealed instantaneously, so we combine confirmed case with Non-Severe, Severe, and Critical cases.

For each person, the state vector can only have one element with value of 1, and the other elements all have value of zero. For example, if a patient is currently in state “Severe Case”, the state vector for him is ‘[0 0 0 1 0 0 0]’. The next day, his state vector could become either ‘[0 0 0 0 1 0 0]’ (Critical Case), ‘[0 0 1 0 0 0 0]’ (Non-Severe Case) or stay the same.

For the sample population, the state vector is defined as the count of people in each state. For example, if there are 100 patients being treated today, out of which 10 are critical, and 90 are severe. The state vector for this sample population is [0 0 0 90 10 0 0]’.

Let’s define the STM as the following:

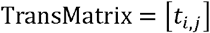

Where

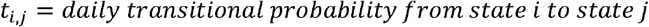

Suppose we have a state vector V(t) for a sample population at time t, how do we predict the state vector V(t+1) in the next day?

Apply simple linear algebra, we can get the following equation:

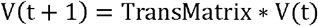

Since the head count of a certain state comes from itself, all other possible transitions into the state (e.g. S has two possible income states, MO and C), minus the outcome states (C, and Cu).

If we want to predict for N period, the equation becomes the following:

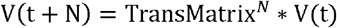

If the population is limited and the transition matrix is stationary, the above formula will be sufficient in predicting all future outcomes. In our case, the population is not fixed, so we need to introduce the additional input into the population: new close contacts (NCC).

Every day, new close contacts are added to the medical observation pool, as people already in the pool will gradually be discharged or confirmed of infection.

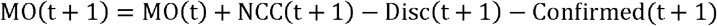

Also, we assume NCC will gradually decay as quarantine measures are put into effect.

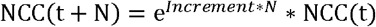

Using this STM model, we will be able predict when the inflection peak time as well as IFP of newly confirmed cases (the maximum open infection cases) in Hubei Province or non-Hubei will occur. Moreover, after verifying this matrix model in China, it could be utilized to evaluate the world epidemic development especially in the major epidemic areas.

Although there is an intermediate state during the above hospitalization: severe cases (the new standard is broken down into mild and normal), critical cases (which can also be divided into general critical and critical), due to the lack of intermediate state transfer probability, we combine the entire hospital period into a in-patient state, for the sake of keeping the model simple. This minimizes the need for only the following five parameters.

- Increment of New Close Contacts (NCC), defined as ln(NCC(t)/NCC(t-1));
- Discharge Rate from Medical Observation (MO), defined as Discharged(t)/MO(t-1)
- Transitional Probability of Medical Observation -> Confirmed cases, defined as Newly confirmed cases (t)/MO(t-1)
- Transitional Probability of Treatment -> Death, defined as New Death Incidents(t) / Treatment(t-1)
- Transitional Probability of Treatment -> Cured, defined as New Cured Incidents (t) / Treatment(t-1)

In order to estimate the count of open non-severe cases, severe cases, and critical cases, we need three more parameters:

- Ratio of Non-Severe Cases
- Ratio of Severe Cases
- Ratio of Critical Cases

### Scenario Setup and Prediction

After validation of the STM model in Hubei Province, we set up three different scenarios derived from China for matching and fitting the major epidemic areas comprising South Korea, Italy, and Iran, in order to control for model error, including optimistic scenario, cautiously optimistic scenario, and relatively pessimistic scenario (Table.1).

**Table.1.** Scenarios for the prediction of outside China. S1: optimistic scenario; S2: cautiously optimistic scenario; S3: relatively pessimistic scenario; Inc: increment.

| **Scenario** | **S1** | **S2** | **S3** |
| --- | --- | --- | --- |
| **Minimum**<br>**Inc** | -30% | -20% | -5% |
| **Daily**<br>**Inc** | -3.87% | -2.20% | -1.50% |

## Results

### The status quo of Hubei Province, China, and the historical prediction model verification of Hubei Province

According to the data of NHC[14], as of Mar 5, there were 67,592 cumulative confirmed cases, 41,966 cumulative cured cases, 126 newly confirmed cases, 29 new deaths, and 1,478 new cured cases, and 19,758 in-patient cases in Hubei Province (Figure.2A). The number of new close contacts in Hubei Province has gradually decreased, and the cumulative number of close contacts is currently 271,959 (Figure.2B). The increment of new close contacts has crossed the IFP (Figure.2C). Based on data from Dec 31, 2019 to Feb 8, 2020 in Hubei Province, we built a prediction model through the STM model, and the cautiously optimistic scenario could consummately predict the arrival of the IFP and several peak dates in Hubei (Table.2), which undoubtedly validate the predictive efficacy of this mathematic model.

**Table.2.** Training set and Validation Set of the Epidemic Trend in Hubei Province. The actual data were extracted from HCHP, and the forecast data in the three scenarios were deduced by the STM model based on the data before Feb 9, 2020. S1: optimistic scenario; S2: cautiously optimistic scenario; S3: relatively pessimistic scenario; MO: medical observation; Inc: increment.

| Key Metrics | Actual | Forecast |  |  |  |  |  |
| --- | --- | --- | --- | --- | --- | --- | --- |
|  |  | S1 |  | S2 |  | S3 |  |
| **Inc of Confirmed Cases** | -9.0% | -10% | -10% | -5% | -5% | -1% | -1% |
| **MO Release Rate** | 16.0% | 17.0% | 10.50% | 17.0% | 10.50% | 17.0% | 10.50% |
| **Peak of Active Cases** | 50,633 | 39,612 | 47,148 | 44,082 | 55,150 | 62,041 | 85,502 |
| **Peak Date** | 2020/2/16 | 2020/2/23 | 2020/2/28 | 2020/3/1 | 2020/3/7 | 2020/4/6 | 2020/4/14 |
| **Peak of Severe Cases** | 9,289 | 5,753 | 6,845 | 6,400 | 8,004 | 9,000 | 12,402 |
| **Peak Date** | 2020/2/16 | 2020/2/23 | 2020/2/28 | 2020/3/1 | 2020/3/7 | 2020/4/6 | 2020/4/14 |
| **Peak of Critical Cases** | 2,492 | 1,786 | 2,124 | 1,986 | 2,484 | 2,793 | 3,849 |
| **Peak Case** | 2020/2/21 | 2020/2/23 | 2020/2/28 | 2020/3/1 | 2020/3/7 | 2020/4/6 | 2020/4/14 |
| **Total Cases at Feb. End** | 66,907 | 54,189 | 64,064 | 60,192 | 71,596 | 68,045 | 81,284 |

**Figure 2.**
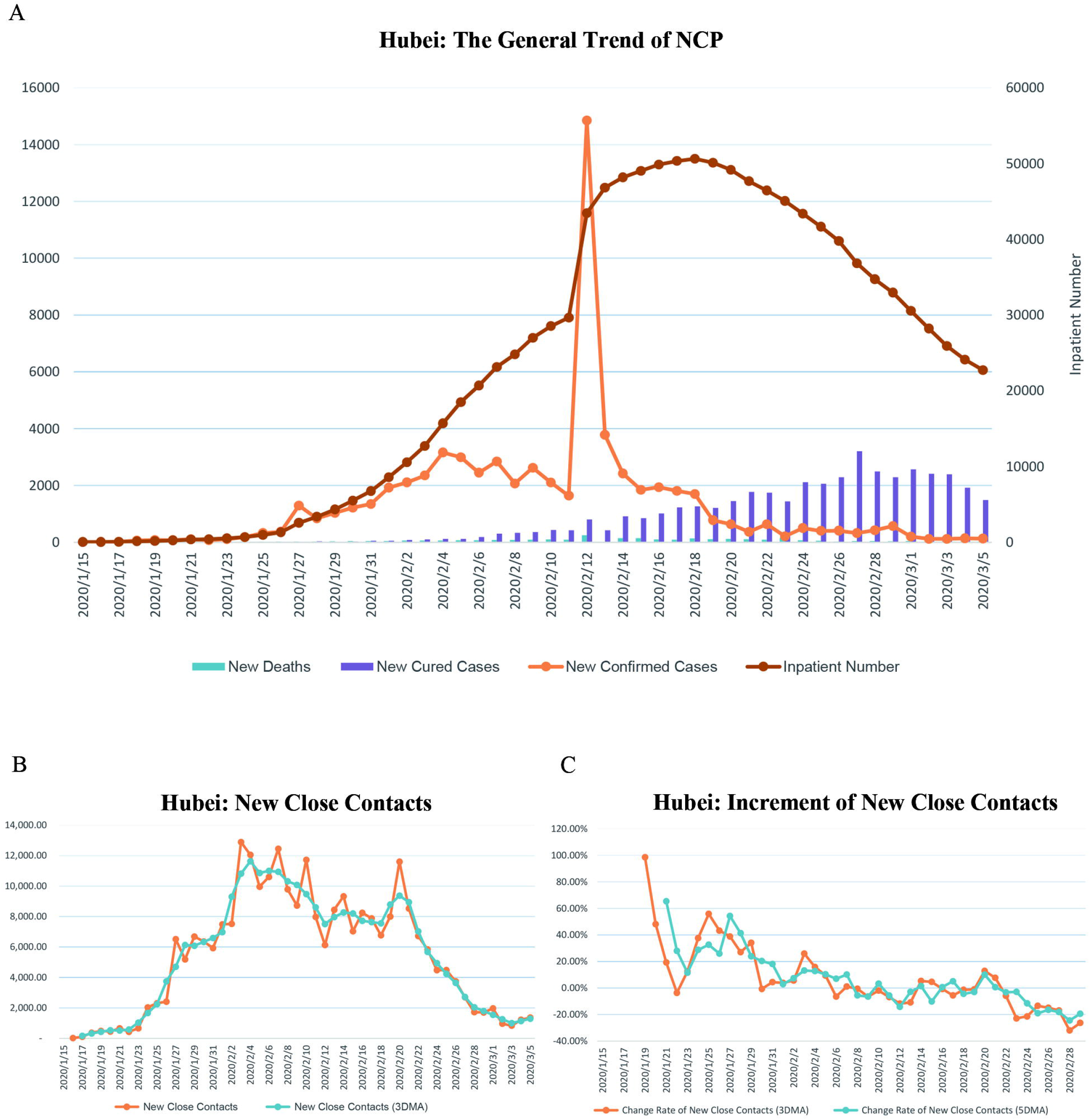
**A**. The epidemic situation and general trend in Hubei Province, including new deaths, new cured cases, newly confirmed cases, and in-patient number from Jan 15, 2020 to Mar 5, 2020. **B**. The trend of new close contacts in Hubei Province from Jan 18, 2020 to Mar 5, 2020. **C**.The increment of new close contacts in Hubei Province from Jan 18, 2020 to Mar 5, 2020. 3DMA: 3-day moving average; 5DMA: 5-day moving average.

### Epidemic situation in training set and the epidemic trend fitting model

As of Mar 5, there were 23,784 confirmed cases, 53,726 cumulative cured cases, 3042 cumulative deaths, 80,552 cumulative confirmed cases, and 670,854 cumulative close contacts in China. Through the analysis, the 5-day moving average (5DMA) and 10-day moving average (10DMA) increment of the confirmed case in Hubei and non-Hubei suggested that the IFP in China was from Feb 6 to Feb 13 (Figure. 3A and B).

**Figure 3.**
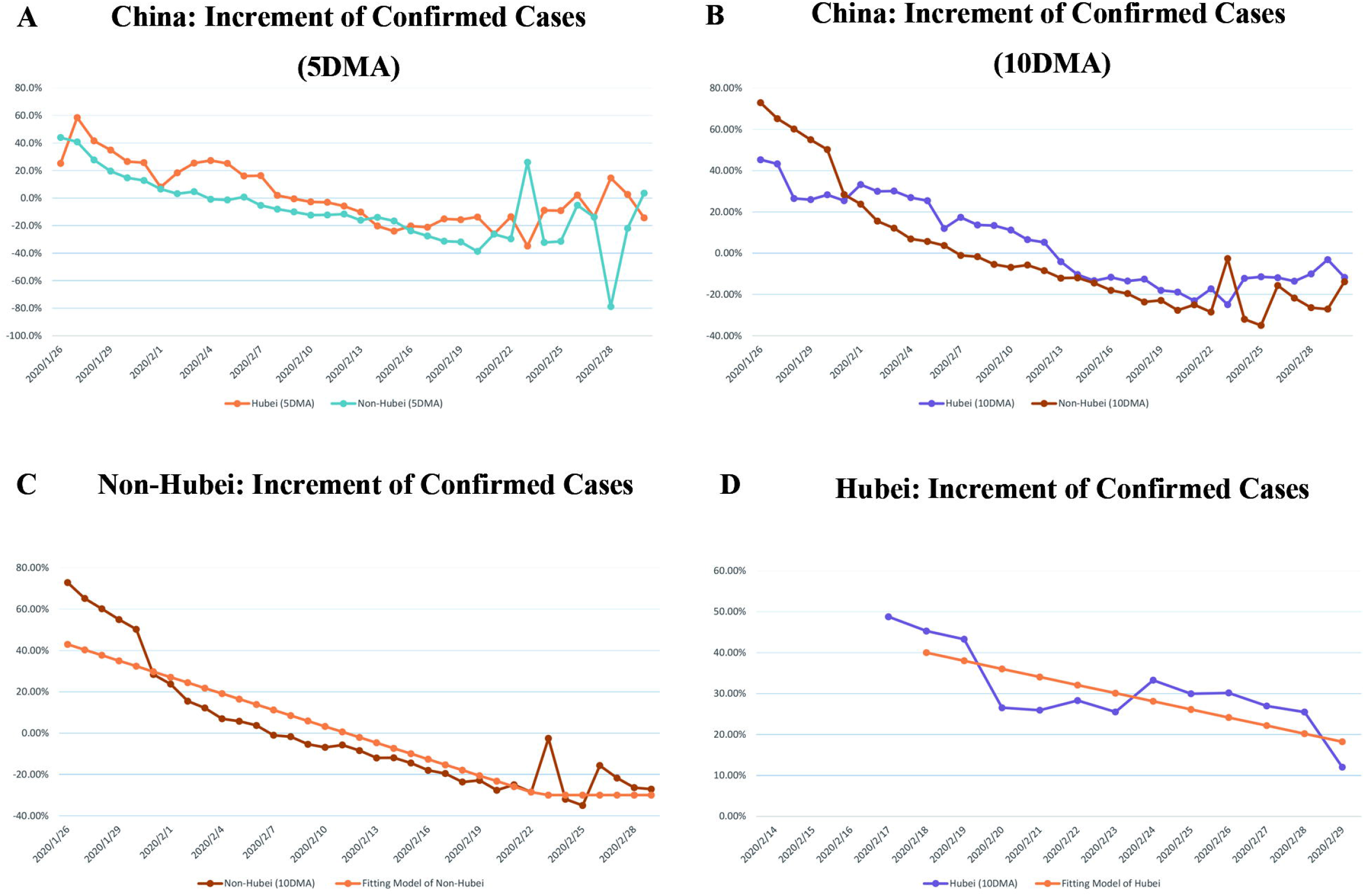
**AB**. The increment of confirmed cases in Hubei and non-Hubei from Jan 22, 2020 to Mar 1, 2020. **C**. The increment and fitting line of confirmed cases in non-Hubei. **D**. The increment and fitting line of confirmed cases in Hubei. 5DMA: 5-day moving average; 10DMA: 10-day moving average.

Applying the STM model again to establish a 10DMA increment of confirmed cases model in non-Hubei, the fitting line of the trend in non-Hubei could be obtained, which is

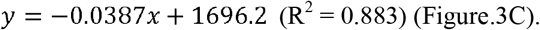

Similarly, in Hubei, the fitting line is

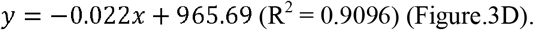

According to the derivatives taken from fitting lines, the epidemic trend in non-Hubei was set as an optimistic scenario with increment of −3.87%, and the epidemic trend in Hubei as a cautiously optimistic scenario with increment of −2.20%, and set a relatively pessimistic scenario with increment of −1.50% (Table.1), which could forecast the situation outside China.

### International epidemic situation and prediction

Data from WHO shows that there were 2,232 new cases worldwide on Mar 5, the cumulative number of confirmed cases reached 95,324, and a total of 85 countries have suffered this epidemic (Figure. 4A) [6]. Starting from the cumulative 50 confirmed cases (T50), the cumulative confirmed case trends were compared in different countries with China, and it showed that the trends of France, Germany, United Kingdom, the United States, and Spain stayed steady, while the trends of newly confirmed cases in Korea, Italy, and Iran laid between Hubei and non-Hubei, which have been identified as the major epidemic areas in the globe (Figure.4B and C).

**Figure 4.**
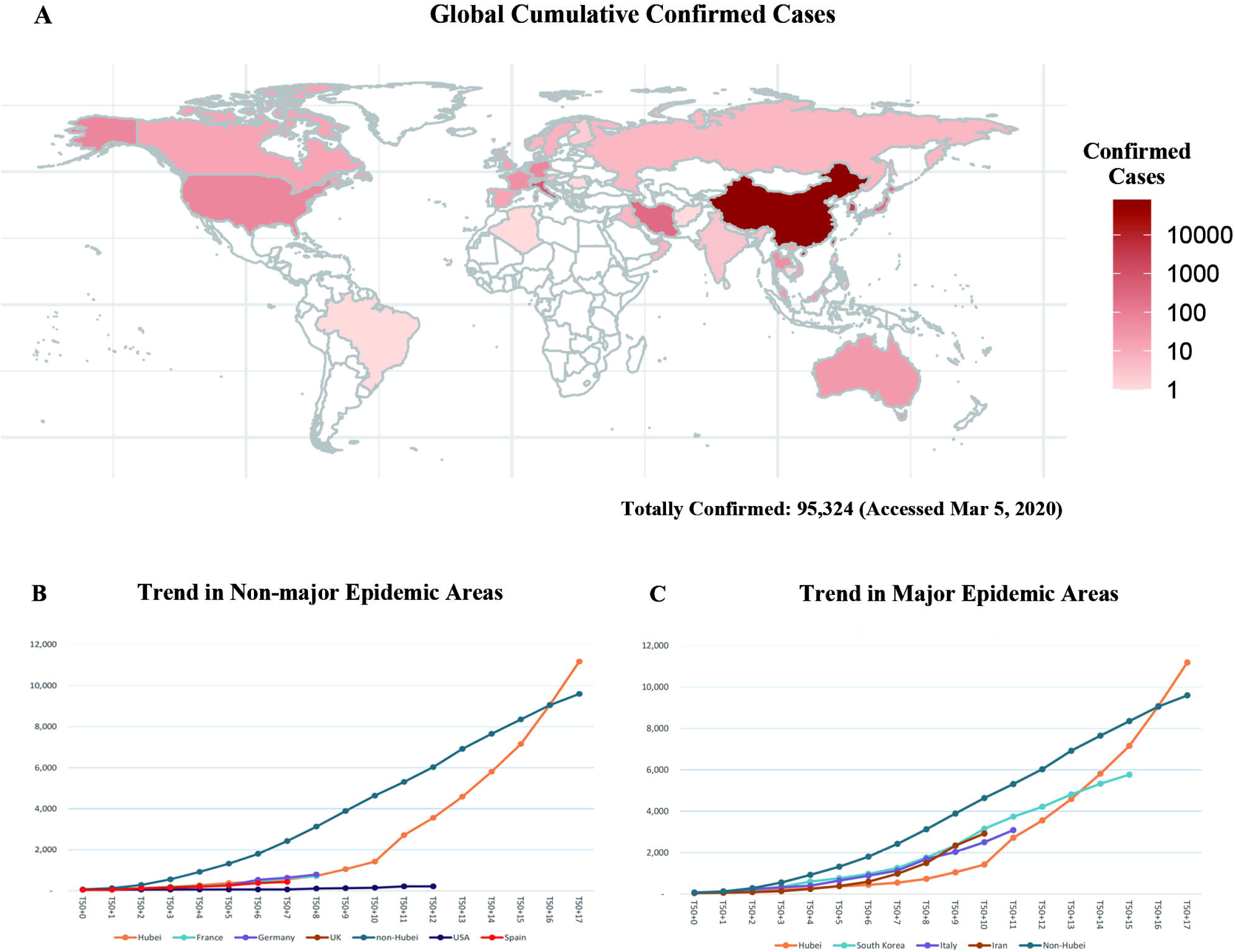
**A**. Global distribution of confirmed cases with totally 95,324 cases on Mar 5, 2020. **BC**. Comparison of the trends in non-major and major epidemic areas.

Then the established STM model was implemented to the three countries. The results showed that the IFP in South Korea would arrive from Mar 6 to Mar 12 (Figure. 5A and B); the IFP in Italy would arrive from Mar 10 to Mar 24 (Figure. 5C and D); the IFP in Iran would come from Mar 10 to Mar 24 (Figure. 5E and F). After completing the model and training set establishment, we compared the cumulative case prediction with the actual data on Mar 6 and Mar 9, which was validation set, and the results overtly testified the efficacy of this prediction model all in Korean, Italy, and Iran (Table.3). By utilizing this model, the approximate number of confirmed cases in the three countries at the end of March, April, and May could be predicted (details show in Figure.6), which could instruct the international medical resources allocation.

**Table.3.** Training Set and Validation Set of the Epidemic Trends in the Major Epidemic Areas. S1: optimistic scenario; S2: cautiously optimistic scenario; S3: relatively pessimistic scenario; Inc: increment.

| Countries | South Korea |  |  |  | Italy |  |  |  | Iran |  |  |  |
| --- | --- | --- | --- | --- | --- | --- | --- | --- | --- | --- | --- | --- |
|  | Actual | Forecast |  |  | Actual | Forecast |  |  | Actual | Forecast |  |  |
| Date |  | S1 | S2 | S3 |  | S1 | S2 | S3 |  | S1 | S2 | S3 |
| 2020/3/5 | 5,766 | 5,766 | 5,766 | 5,766 | 3,089 | 3,089 | 3,089 | 3,089 | 2,922 | 2,922 | 2,922 | 2,922 |
| 2020/3/6 | 6,284 | 6,199 | 6,221 | 6,239 | 3,858 | 3,794 | 3,844 | 3,852 | 3,513 | 3,613 | 3,678 | 3,684 |
| 2020/3/7 | 6,767 | 6,612 | 6,683 | 6,741 | 4,636 | 4,609 | 4,795 | 4,830 | 4,747 | 4,396 | 4,630 | 4,671 |
| 2020/3/8 | 7,134 | 6,990 | 7,142 | 7,268 | 5,883 | 5,515 | 5,965 | 6,064 | 5,823 | 5,250 | 5,806 | 5,921 |
| 2020/3/9 | 7,382 | 7,323 | 7,589 | 7,811 | 7,375 | 6,485 | 7,374 | 7,598 | 6,566 | 6,147 | 7,224 | 7,481 |

**Table 4.**
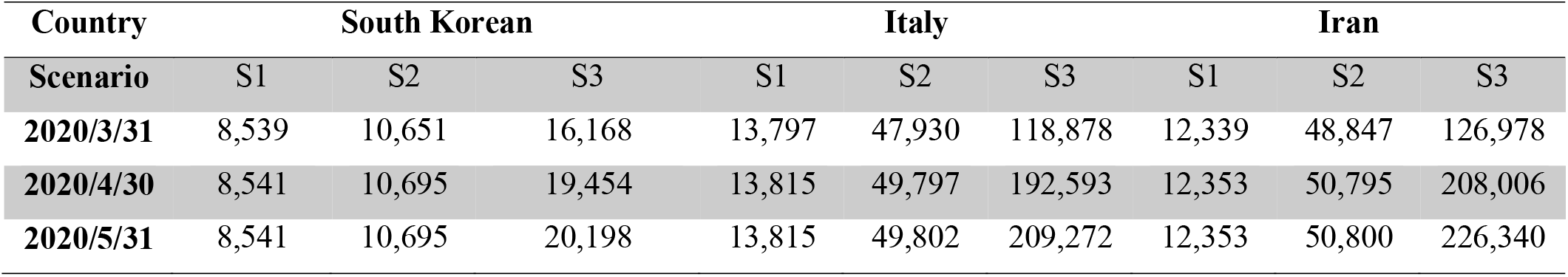
Predictive Cumulative Confirmed Cases in the Major Epidemic Areas. S1: optimistic scenario; S2: cautiously optimistic scenario; S3: relatively pessimistic scenario.

| Country | South Korean |  |  | Italy |  |  | Iran |  |  |
| --- | --- | --- | --- | --- | --- | --- | --- | --- | --- |
| Scenario | S1 | S2 | S3 | S1 | S2 | S3 | S1 | S2 | S3 |
| 2020/3/31 | 8,539 | 10,651 | 16,168 | 13,797 | 47,930 | 118,878 | 12,339 | 48,847 | 126,978 |
| 2020/4/30 | 8,541 | 10,695 | 19,454 | 13,815 | 49,797 | 192,593 | 12,353 | 50,795 | 208,006 |
| 2020/5/31 | 8,541 | 10,695 | 20,198 | 13,815 | 49,802 | 209,272 | 12,353 | 50,800 | 226,340 |

**Figure 5.**
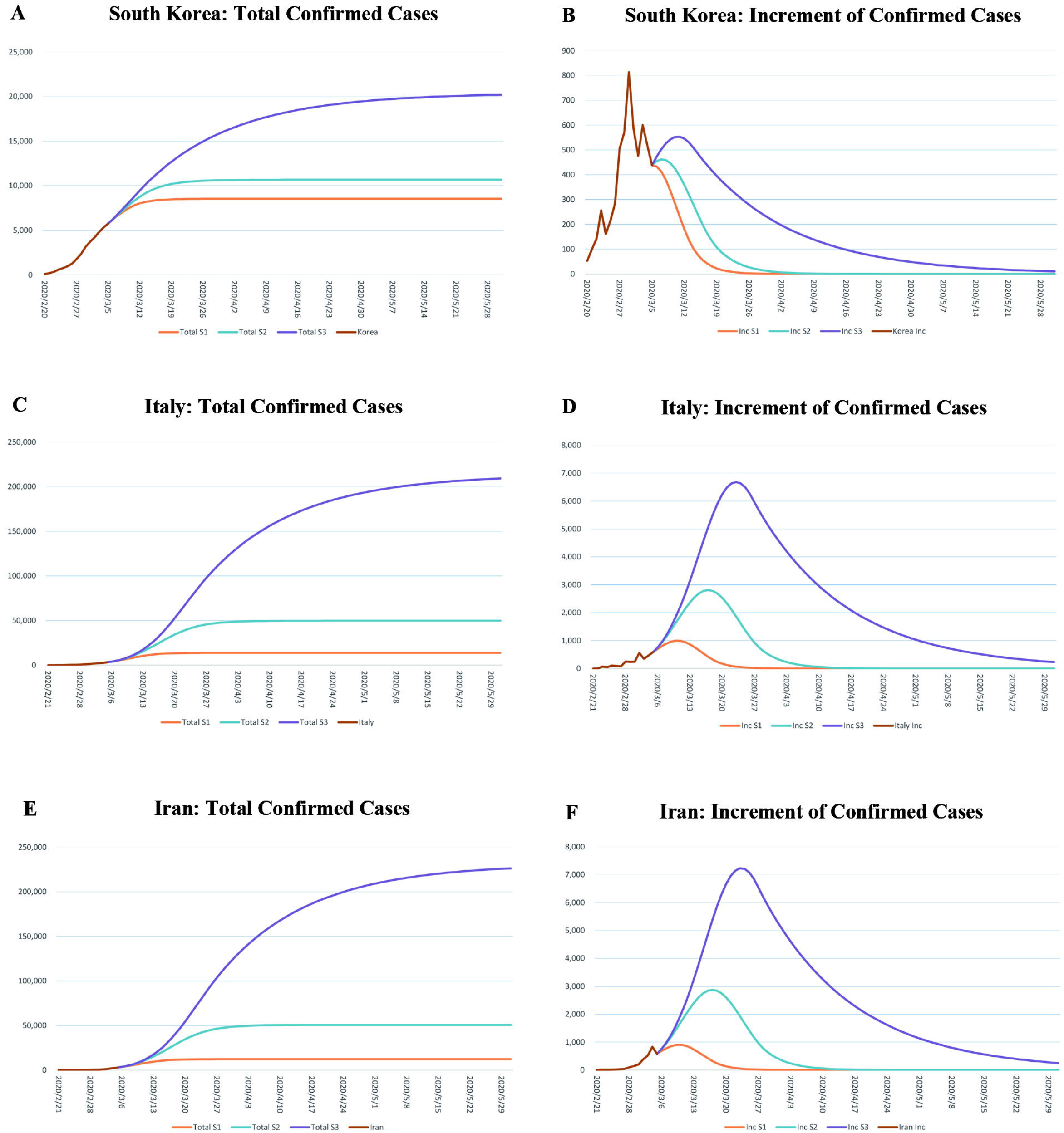
**A to F**. Predictive total confirmed cases and increment of confirmed cases in South Korean, Italy, and Iran, respectively, with the three scenarios deduced by the State Transition Matrix Model based on the data before Mar 6, 2020.

**Figure 6.**
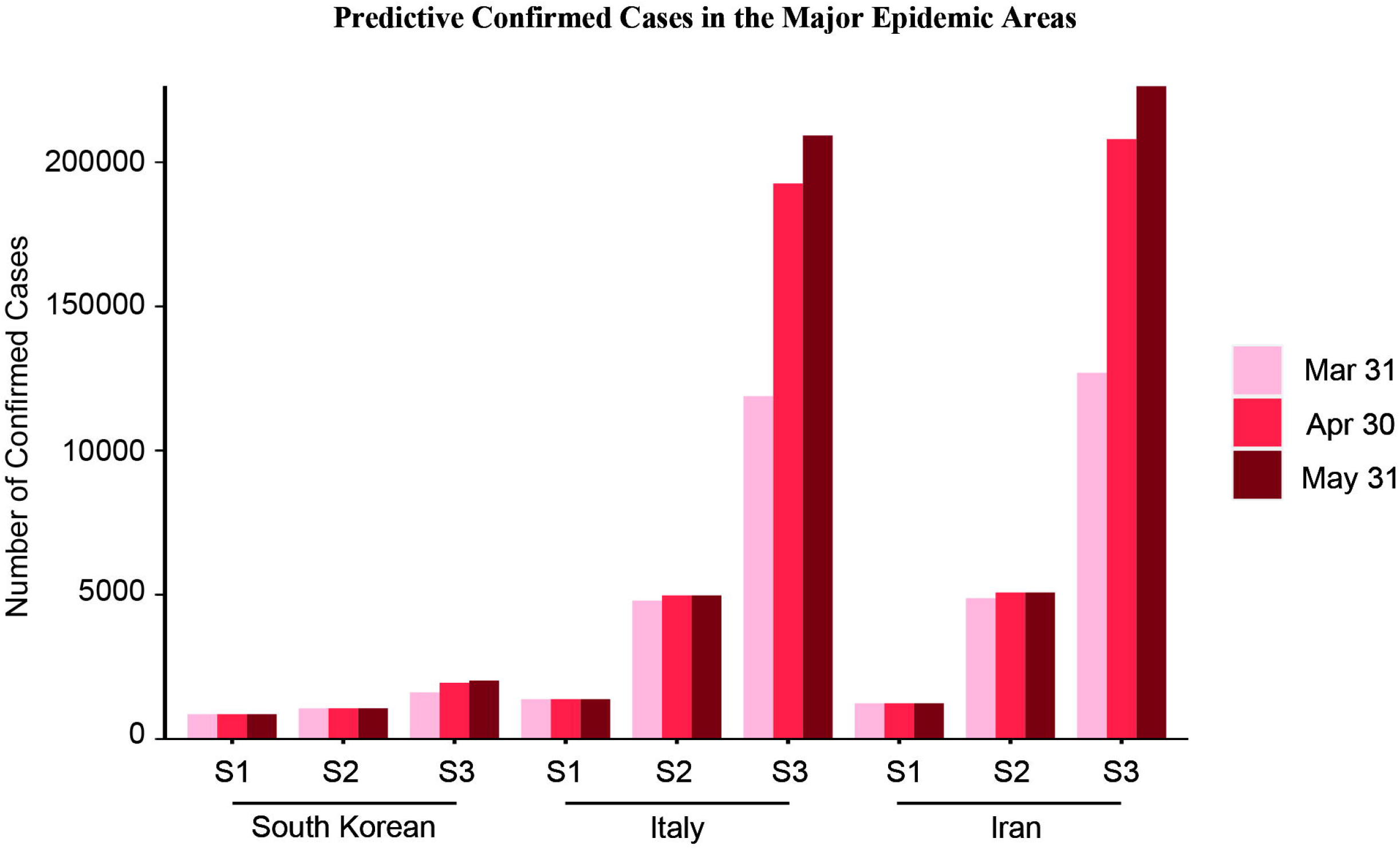
Predictive cumulative confirmed cases in South Korean, Italy, and Iran, respectively, on Mar 31, Apr 30, and May 31. See also Table.4.

## Discussion

Given the punchy transmissibility of COVID-19 [15], isolation and quarantine are undoubtedly the primary options [11]. Subsequently, the model of this epidemic sprouted out a lot. Ziff *et al*. established a model of death cases and reported that death cases follow three patterns: exponential growth, power-law behavior, and then exponential decline in the daily rate [16]. Nevertheless, deaths are affected by many factors, such as age [17, 18]. More attention should be paid to the number of new cases, and the rate of increment, attributed to the effect of epidemic prevention and control, can be evaluated to guide the date of return to work.

Moreover, we must strictly follow the coping strategy and learn the Chinese model for dealing with NCP outbreaks. Li *et al*. developed a simple regression model, and based on this model, they estimated that about 34 founder patients outside of China were not observed in the early stage of transmission, and the global trend approximated an exponential increase, tenfold increase in 19 days [19]. This study reproduced the disease’s initial spread to the world, yet made no prediction for the future trend, and exponential growth will be curbed immediately after the attention of local governments, and the IFP will come. Milan Batista proposed an estimate of the final size of the COVID-19 epidemic, the logistic growth model and classic susceptible-infected-recovered dynamic model are used to estimate the final size of the coronavirus epidemic, being approximately 83700 (±1300) cases and that the peak of the epidemic was on Feb 9 2020 [20]. However, as of Mar 5, the number of global cases has reached 95,333, and the IFP for growth in South Korea, Italy, and Iran has not yet arrived, which means the global size will be even more colossal.

Our model is based on the fitting of real data from authorities. Through the STM Model, based on data from Hubei and non-Hubei, we predict the IFPs in Korea, Italy, and Iran, while there are still some limitations. Due to the large outbreaks started at different times all over the world, the effects of seasonal and geographical factors have not been taken into account. Although the fitting with the Chinese model can better predict the situation around the world, through reference and learning, the response strategies of other countries may be more mature. As China resumes work, the production capacity of various medical resources will gear up rapidly, which will impose a positive impact on the world, and it could be more optimistic that the IFP will come soon.

Local governments, regardless of the speed of outbreaks, should learn from China’s primary response strategy, such as stopping working, reducing gathering, preventing contact transmission, wearing masks, and implementing quarantine. After the NCP being under control, the production and output of medical resources should be intensified, the production of coronavirus detection kits should be accelerated, existing cases should be summarized. More accurate diagnostic criteria should be compiled to prevent massive missed diagnoses in countries lacking the kit. Even if it currently causes some global economic regression, the recovery will swiftly come after holding the throat of NCP and COVID-19.

## Conclusion

Based on data from China, we utilized the State Transition Matrix Model to predict the IFP of disease in countries currently experiencing outbreaks worldwide. If properly controlled, the IFP in South Korea and Italy will come in early March, and the IFP in Iran will come in mid-March. During this period, countries around the world should work together to fight the epidemic.

## Data Availability

The data that support the findings of this study are available in the National Health Commission of the People*s Republic of China, the Health Commission of Hubei Province, and the World Health Organization.

http://www.nhc.gov.cn/

http://wjw.hubei.gov.cn/

https://www.who.int/emergencies/diseases/novel-coronavirus-2019/situation-reports/

## Acknowledgements

Not applicable.

## Authors’ contributions

Jian Chen and Junhua Zheng participated in study design; Zhong Zheng and Ke Wu performed data collection and analysis; Zhixian Yao drafted the manuscript; all author provided critical review of the manuscript and approved the final draft for publication.

